# Integrative analyses identify potential key genes and pathways in Keshan disease using whole-exome sequencing

**DOI:** 10.1101/2021.03.12.21253491

**Authors:** Jichang Huang, Chenqing Zheng, Rong Luo, Mingjiang Liu, Qingquan Gu, Jinshu Li, Xiushan Wu, Zhenglin Yang, Xia Shen, Xiaoping Li

**Affiliations:** Institute of Geriatric Cardiovascular Disease, Chengdu Medical College, Chengdu, People’s Republic of China; State Key Laboratory of Biocontrol, School of Life Sciences, Sun Yat-sen University, Guangzhou, China; Department of Cardiology, Hospital of the University of Electronic Science and Technology of China and Sichuan Provincial People’s Hospital, Chengdu, Sichuan, China; Shenzhen RealOmics (Biotech) Co., Ltd., Shenzhen, China; Institute of Endemic Disease, Center for Disease Control and Prevention of Sichuan Province, Chengdu, Sichuan, China; The Center of Heart Development, College of Life Sciences, Hunan Norma University, Changsha, China

**Keywords:** Keshan disease, dilated cardiomyopathy, whole-exome sequencing, gene-based burden analysis, automated Phenolyzer analysis

## Abstract

Keshan disease (KD), an endemic heart disease with multifocal necrosis and replacement fibrosis of the myocardium,is still a nightmare situation for human health. However, molecular mechanism in the pathogenesis of KD remains unclear. Herein, blood samples were collected from 68 KD patients and 100 controls, and we systematically analyzed mutation profiles using whole-exome sequencing (WES). Causative genes of dilated cardiomyopathy (DCM), gene-based burden analysis, disease and pathway enrichment analysis, and protein-protein interaction (PPI) network analysis were performed. Of the 98 DCM-causative genes, 106 rare variants in 28 genes were detected in KD patients with minor allele frequency (MAF) < 0.001. Gene-based burden analysis, PPI network analysis, and automated Phenolyzer analysis were performed to prioritize 199 candidate genes, which combined with 98 DCM-causative genes, and reference genes from gene microarray or proteomics in KD. Then, 19 candidate pathogenic genes were selected, and 9 candidate genes were identified using PPI analysis, including *HIF1A, GART, ALAD, VCL, DTNA, NEXN, INPPL1, NOS3*, and *JAK2*. The 199 candidate genes were further analyzed using disease enrichment with CTD database and PPI analysis, and 21 candidate genes were identified. By combining with disease enrichment and PPI analysis, 7 Selenium (Se)-related genes were further identified, including *ALAD, RBM10, GSN, GGT1, ADD1, PARP1*, and *NOS3*. Based on the gene function and data validation, *NEXN, TAF1C, FUT4, ALAD, ZNF608*, and *STX2* were the most likely pathogenic genes in KD. Notably, *ALAD* is the only candidate pathogenic gene identified by four different analyses, and its homozygous mutant mice could affect heart development and cause death.

## 1. Introduction

Keshan disease (KD), a dilated cardiomyopathy, still has a high mortality. KD was firstly noted in 1935 in Keshan County, Northeast China [1], whilst it was reported in Nagano Prefecture in Japan and northern mountains in North Korea in the 1950s as well [2]. It mainly invades the myocardium, manifested as myocardial degeneration, chronic heart failure, various arrhythmias and even sudden death [3]. The effective treatments used in Keshan disease are not available till now.

Selenium deficiency is believed as the direct cause for KD occurrence [4]. Selenium supplementation for the treatment of Keshan disease has been shown significant clinical benefits, while it is difficult to explain the seasonal or annual variation of KD occurrence [5, 6]. Moreover, KD disease also occurs in some regions where selenium is not lacking [7]. Indeed, there is increasing evidence that Keshan disease is related to polymorphisms, familial clustering, and mutations of related genes, indicating that genetic factors are important promoters for the etiology and molecular mechanism of this disease [8]. To explore the possible mechanism, genetic analysis should be performed.

Recently, owing to the advancement in whole exome sequencing (WES), it has been successfully used to discover disease-related genes of for multiple Mendelian or non-Mendelian diseases [9]. However, Genome-Wide Association Studies (GWAS) focuses on common mutations (MAF ≥ 5%), and its impact is small, RR≈1.2-1.5 [10]. In addition, the large number of GWAS SNPs explains a small part of the heritability of the disease [10]. Compared to common variant studies, individual SNP analysis in rare variant studies is seriously underpowered [11]. Recent advances in gene-based burden analysis might help to tackle some of these limitations. The approach was based on gene-based burden analysis, in which the total frequency of “qualifying variants” of each gene is compared between cases and controls [11]. Therefore, we hypothesize that the application of this more integrated approach may help elucidate the genetic etiology of Keshan disease.

As far s we know, the pathogenic gene of Keshan disease has not been reported. In this study, we approached WES to identify potential key genes by gene-based burden analysis, KEGG pathway enrichment, and protein-protein interaction (PPI) analyses for KD disease.

## 2. Results

### 2.1 Clinical baseline data of KD patients

The subjects, including 68 KD patients and 100 healthy individuals, were recruited into this study. Of 68 KD patients, it is defined as left ventricle enlargement (men > 55mm, women > 50mm) and reduced ejection fraction (LVEF < 50%) obtained by echocardiography. Among the KD patients, 30 females (44.1%, 30/68) and 38 males (56.7%, 38/68), a mean age 56.81 ± 11.01 years old, were included in our study. As shown in Table 1, the percentage of patients with NYHA class II/III, left bundle-branch block (LBBB), right bundle-branch block (RBBB), intraventricular conduction delay, ventricular premature beats, atrial fibrillation were 83.8% (57/68), 7.4% (5/68), 14.7% (10/68), 26.5% (18/68), 22.1% (15/68), 13.2% (9/68), respectively. The left ventricular diameter was 55.33 ±10.97 mm.

**Table 1.** Demographic baseline of patients

| Variables | Total patients (n = 68) |
| --- | --- |
| Sex, male (%) | 38 (56.7) |
| Age, year | $56.81 \pm 11.01$ |
| BMI, kg/m<sup>2</sup> | $57.24 \pm 10.83$ |
| Smoking (%) | 16 (23.5) |
| Drinking (%) | 16 (23.5) |
| NYHA II/III (%) | 57 (83.8) |
| History of cardiovascular disease (%) | 28 (41.2) |
| The left atrium diameter (mm) | $38.24 \pm 8.52$ |
| The left ventricular diameter (mm) | $55.33 \pm 10.97$ |
| The right ventricular diameter (mm) | $22.00 \pm 3.20$ |
| LBBB (%) | 5 (7.4) |
| RBBB (%) | 10 (14.7) |
| Intraventricular conduction delay (%) | 18 (26.5) |
| Ventricular premature beats (%) | 15 (22.1) |
| Atrial fibrillation (%) | 9 (13.2) |

### 2.2 Rare variants of DCM-causative genes in KD patients

To elucidate the possible genetic mechanisms of KD pathogenesis, a dilated cardiomyopathy, rare variants of DCM-causative genes were measured in accordance with minor allele frequency (MAF) below 0.1% in the ExAC and the database for the 1000 Genomes Project [12,13]. There are various rare variants of DCM-causative genes in KD patients as follows: 34 rare variants in *TTN* (n = 34), 20 in *OBSCN* (n = 20), 4 in *LAMA4* (n = 4), 2 in *MYBPC3* (n = 2), 2 in *VCL* (n = 2), 3 in *FHL2* (n = 3), 3 in *ZBTB17* (n = 3), 2 in *RBM20* (n = 2), 2 in *ANKRD1* (n = 2), 2 in *KCNQ1* (n = 2), 4 in *DMD* (n = 4), 2 in *MYPN* (n = 2), x in *MYH7* (n = 2), 2 in *BRAF* (n = 2), 4 in *FLNC* (n = 4), 1 in *MYH6* (n = 1), 3 in *SYNM* (n = 3), 2 in *RYR2* (n = 2), 1 in *ABCC9* (n = 1), 1 in *DSC2* (n = 1), 1 in *FKRP* (n = 1), 1 in *FKTN* (n = 1), 2 in *LDB3* (n = 2), 2 in *SCN5A* (n = 2), 1 in *TXNRD2* (n = 1), 1 in *SOS1* (n = 1), 1 in *MURC* (n = 1), 1 in *DSG2* (n = 1) as shown in Supplementary Table S2 and Figure 1. These genes were associated with various biological processes, including sarcomere, cytoskeleton, desmosomes, sarcoplasmic reticulum, nucleus, ion channel, extracellular matrix, and mitochondria.

**Figure 1.**
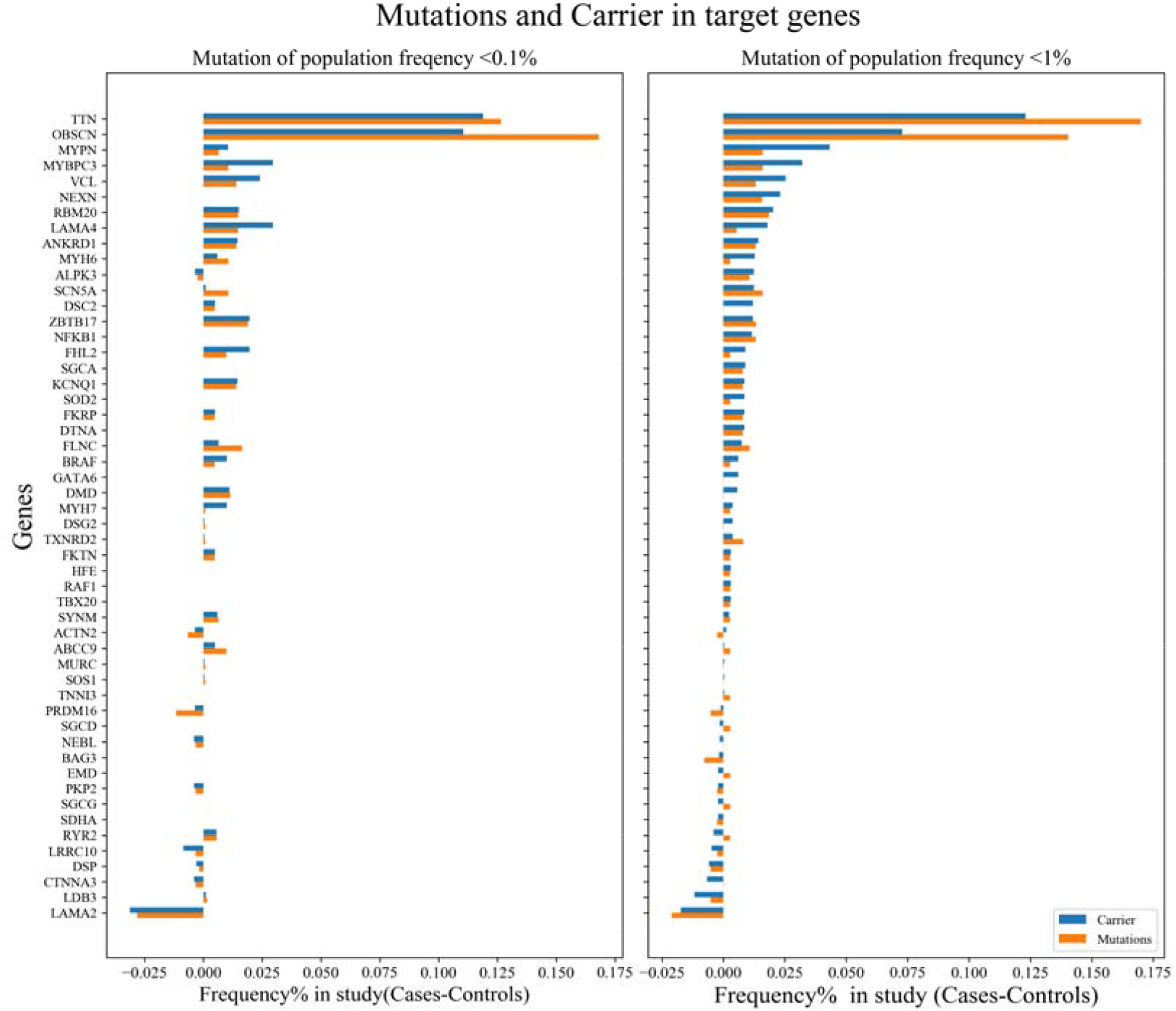
The number of rare variants and cases in DCM-causative genes with MAF < 0.001(A) and MAF < 0.01 (B). The blue column shows the number of rare variants of DCM-causative genes, and the yellow column show the number of patients with rare variants.

As the incidence of KD was dramatically reduced and a relatively small sample sizes were selected in this study, so we also have applied rare variants with MAF below 1% to further analyze. In total, the 253 rare variants were identified in 52 genes as shown in Figure 1. These genes were related to sarcomere, cytoskeleton, desmosomes, sarcoplasmic reticulum, nucleus, ion channel, extracellular matrix, and mitochondria. These results were similar to rare variants with MAF below 0.1%, further suggesting that these biological processes may play an important role in KD pathogenesis.

### 2.3 Gene-based burden analysis and identification of candidate genes

To further reveal the aggregate association of rare variants, gene-based burden analysis were introduced to obtain a gene-level mutational profile (MAF < 0.001, MAF < 0.01 or MAF < 0.05; *p*-value < 0.05). Manhattan figures and Q-Q plots of gene-based GWAS results were displayed in Supplemental Figure 1. Rare variants of 199 genes were listed in Table S3. Among these genes, five DCM-causative genes, including *MYBPC3, OBSCN, DTNA, NEXN*, and *VCL* were identified in KD patients.

To select the potential pathogenic genes from 199 genes, comprehensive protein–protein interactions (PPI) were proposed to prioritize candidate genes, which combined with 98 DCM-causative genes, reference genes from gene microarray in KD, and reference genes from proteomics in KD [14]. Top 30 genes were selected to constitute gene sets from DCM-causative genes, gene microarray, and proteomics, respectively. Then, a Venn diagram was performed for intersect genes selection from three-gene sets. There were 3 genes in all three-gene sets in Figure 2A, including *HIF1A, GART*, and *ALAD*. 16 intersect genes were screened in any two-gene sets: *MYOM1, VCL, DTNA, KIAA1217, RBMX, NEXN, MYOT, ABCB5, LUZP1, JAK2, ERAL1, INPPL1, ETV4, NOS3, RICTOR*, and *LDHAL6B*. Notably, *VCL, DTNA, and NEXN* were identified in DCM as causative genes.

**Figure 2.**
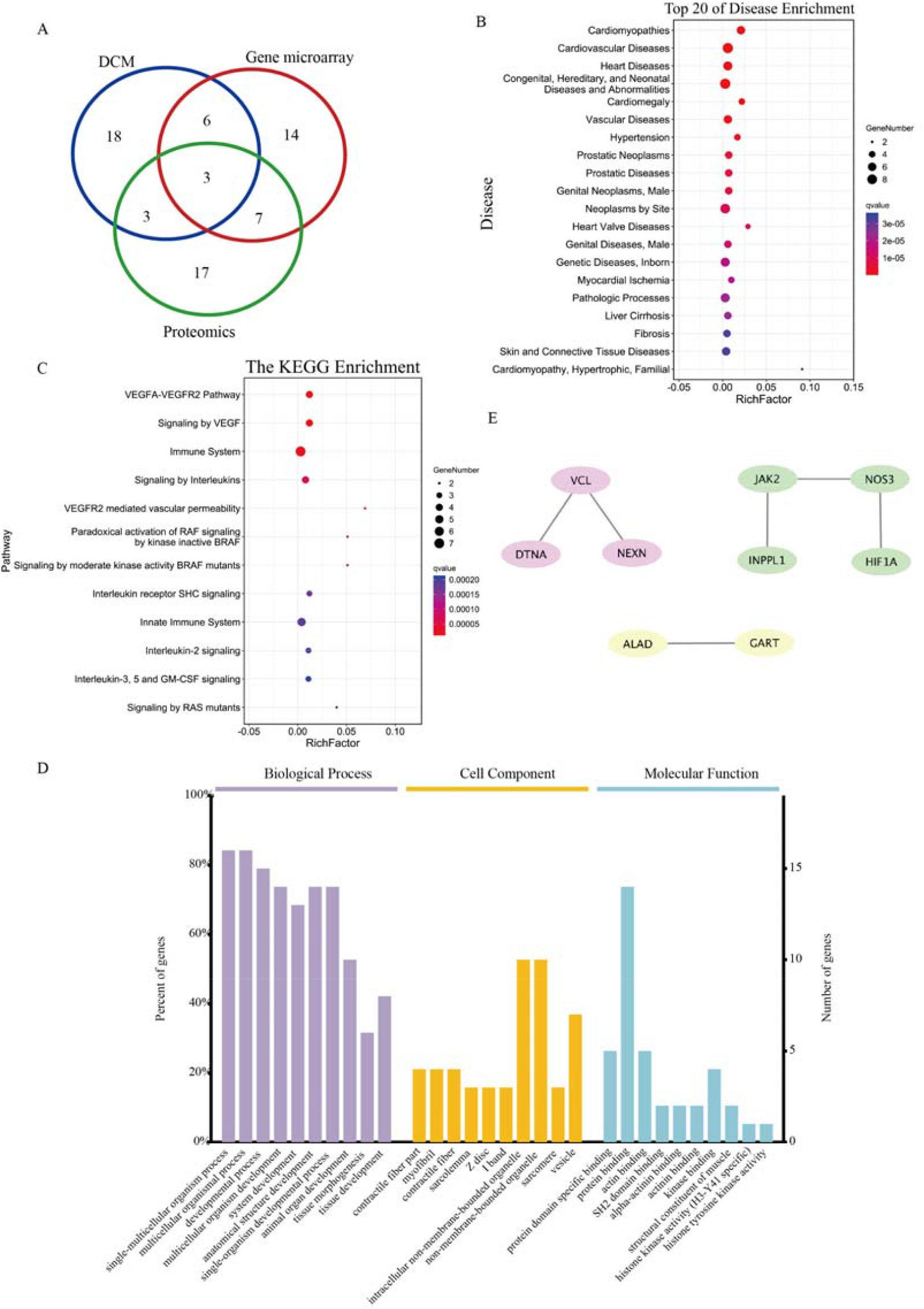
Identification and functional annotation of candidate genes. (A) The overlap of genes from burden analysis in DCM-causative genes, gene microarray, and proteomics. (B) The bubble chart of top

The disease enrichment analysis was conducted to identify the relationship between diseases and these 19 genes through the Comparative Toxicgenomics Database (CTD) database. The top 20 enriched diseases are shown in Figure 2B. Of the top 20 diseases, 10 diseases might be closely related with KD, including 1 fibrosis, and 9 cardiovascular diseases. 20 diseases with 18 overlapping genes enriched by the Comparative Toxicgenomics Database (CTD) database. (C) The bubble chart of top 20 pathways with eighteen overlapping genes enriched by Reactome database. (D) The Gene Ontology (GO) analysis of eighteen overlapping genes. (E) The protein–protein interactions (PPI) analysis of eighteen overlapping genes.

The *Kyoto Encyclopedia of Genes and Genomes* (KEGG) pathway enrichment analysis revealed that 18 candidate genes might be associated with signaling by VEGF, immune system, signaling by interleukins, paradoxical activation of RAF signaling, and MAPK family signaling cascades (Figure 2C). In addition, the Gene Ontology (GO) analysis revealed that these genes might be related to contractile fiber, myofibril, sarcolemma, Z disc, I band, vesicle, sarcomere, and actin binding (Figure 2D). The protein–protein interactions (PPI) analysis was performed to further screen the most likely candidate genes, and there were 9 genes constituting PPI network, including *HIF1A, GART, ALAD, VCL, DTNA, NEXN, INPPL1, NOS3*, and *JAK2* (Figure 2E).

Based on the above-related analysis and the previous functional study, the candidate genes (*HIF1A, GART, ALAD, VCL, DTNA, NEXN, INPPL1, NOS3*, and *JAK2*) were further considered as the most likely genes.

### 2.4 Enriched disease analysis and functional annotation of candidate genes

The disease enrichment analysis was conducted to identify the relationship between diseases and these genes through the CTD database. A total of 59 different diseases were significantly enriched by the genes with an MAF < 0.001, MAF < 0.01, and MAF < 0.05, respectively. The top 30 enriched diseases are shown in Figure 4. Of the top 30 diseases, 5 cardiovascular diseases might be closely related with KD. Subsequently, except for the top 30 enriched diseases, there were 7 cardiovascular diseases.

**Figure 3.**
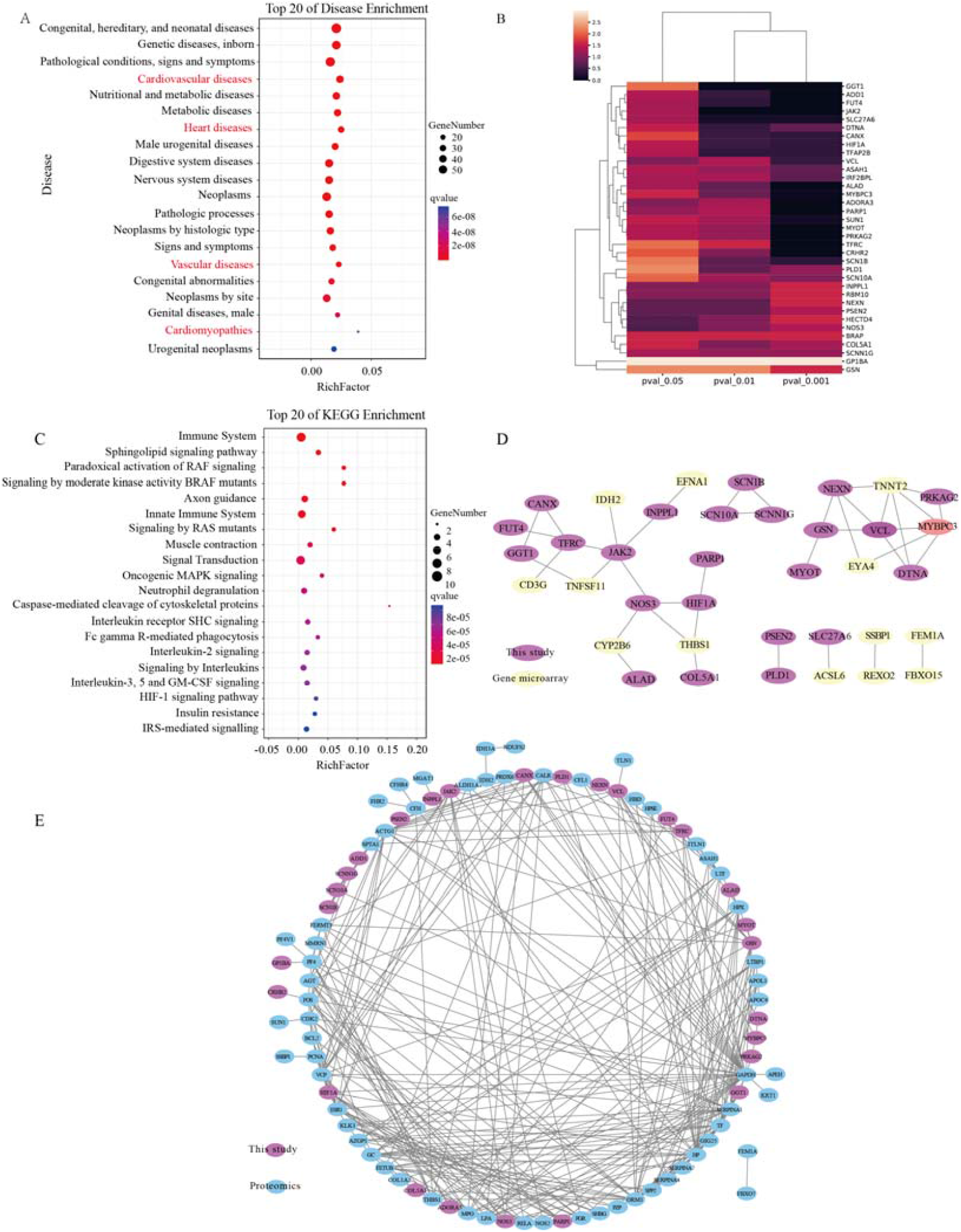
Enriched disease analysis and functional annotation of candidate genes. (A) The bubble chart of top 20 diseases with 199 genes from burden analysis enriched by the CTD database. (B) The mutation profile of 35 candidate genes. (C) The bubble chart of top 20 pathways with eighteen overlapping genes enriched by Reactome database. (D) The PPI analysis of 35 candidate genes, genes from gene microarray. Purple, and yellow circles represent genes from our study and gene microarray in previous study, respectively. (E) The PPI analysis of 35 candidate genes and genes from proteomics. Purple and blue circles represent genes from our study and proteomics in previous study, respectively.

**Figure 4.**
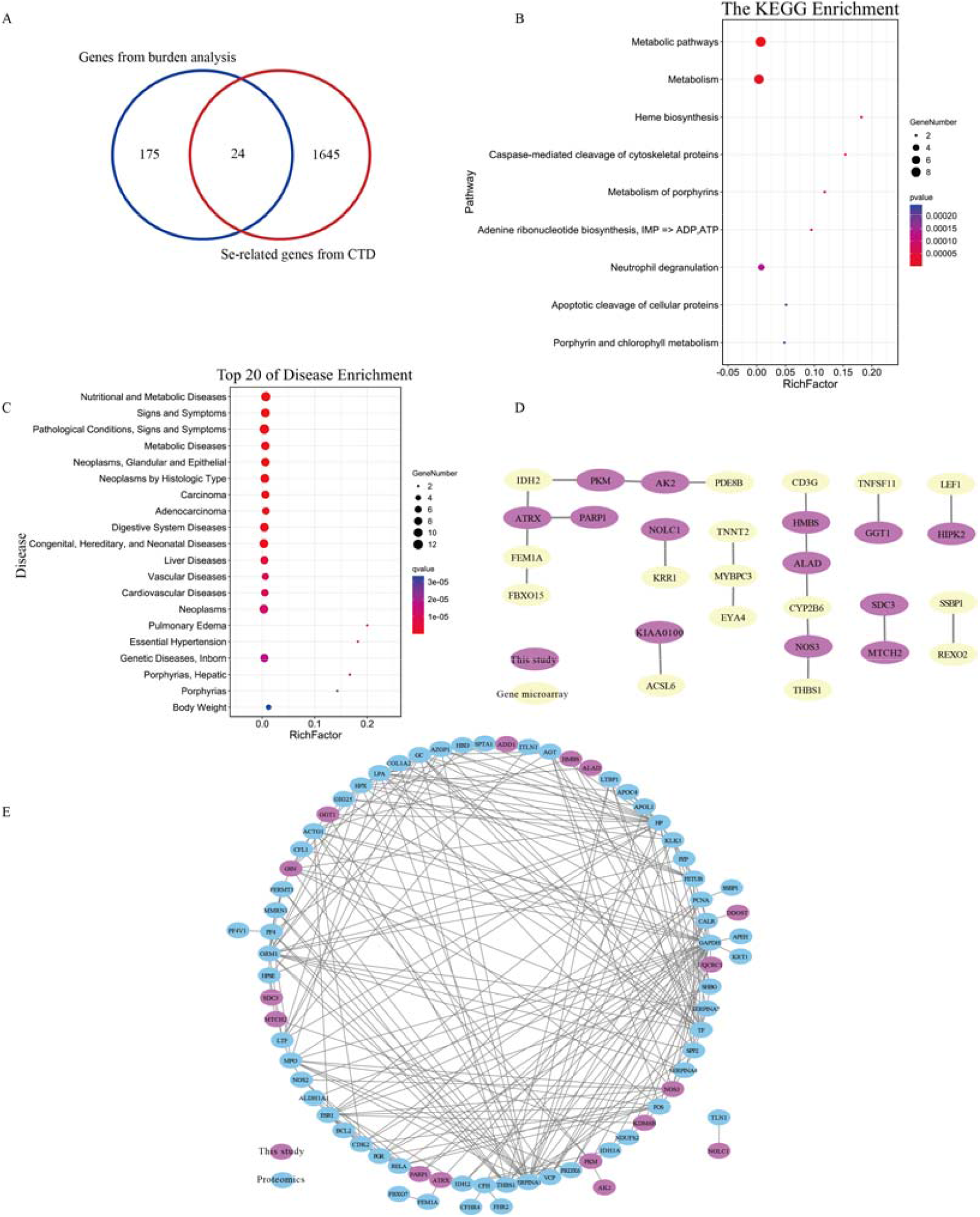
Identification and functional annotation of Se-related genes. (A) The overlap of genes from burden analysis and Se-related genes from CTD database. (B) The bubble chart of top 20 pathways with 24 overlapping genes enriched by Reactome database. (C) The bubble chart of top 20 diseases with 24 genes enriched by CTD database. (D) The PPI analysis of 24 Se-related genes and genes from gene microarray. Purple and yellow circles represent genes from our study and gene microarray in previous study, respectively. (E) The PPI analysis of 24 Se-related genes and genes from proteomics. Purple and blue circles represent genes from our study and proteomics in previous study, respectively.

Based on the analysis given above, we identified 35 candidate pathogenic genes: *ADD1, ADORA3, ALAD, ASAH1, BRAP, CANX, COL5A1, CRHR2, DTNA, FUT4, GGT1, GP1BA, GSN, HECTD4, HIF1A, INPPL1, IRF2BPL, JAK2, MYBPC3, MYOT, NEXN, NOS3, PARP1, PLD1, PRKAG2, PSEN2, RBM10, SCN10A, SCN1B, SCNN1G*,0020*SLC27A6, SUN1, TFAP2B, TFRC*, and *VCL* (Figure 3A). The mutation profile of 35 genes was shown in heat map with an MAF < 0.001, MAF < 0.01, and MAF < 0.05 (Figure 3B). Notably, 4 DCM-causative genes, including *MYBPC3, DTNA*, NEXN, and VCL were discovered in KD patients. These 35 genes were associated with various biological processes, including immune system, RAF signaling pathway, axon guidance, signaling by interleukins, HIF-1 signaling pathway, MAPK signaling, and IRS-mediated signaling (Figure 3C).

To further elucidate the pathogenic genes from 35 candidate genes, PPI analysis were proposed to prioritize candidate disease genes, which combined with genes from microarray in KD, and genes from proteomics in KD. There were 21 genes constituting 4 networks, including network 1 (*SCN10A, SCN1B*, and *SCNN1G*), network 2 (*MYOT, GSN, NEXN, VCL, DTNA, MYBPC3*, and *PRKAG2)*, network 3 (*GGT1, CANX, TFRC, FUT4, JAK2, INPPL1, NOS3, HIF1A*, and *PARP1*), network 4 (*PSEN2*, and *PLD1*) in Figure 3D.

The PPI analysis was further proposed to prioritize candidate disease genes, which combined with genes from proteomics in KD. There were 27 genes, including *ADD1, ADORA3, ALAD, CANX, COL5A1, CRHR2, DTNA, FUT4, GGT1, GP1BA, GSN, HIF1A, INPPL1, JAK2, MYBPC3, MYOT, NEXN, NOS3, PARP1, PLD1, PRKAG2, PSEN2, SCN10A, SCN1B, SCNN1G, TFRC*, and *VCL* in Figure 3E.

### 2.5 Screening of the Se-related genes in gene-based burden results

Because Se deficiency was closely connected with KD, we further identified 24 candidate Se-related genes from gene-based burden results through the CTD database: *ADD1, AK2, ALAD, ATRX, DDOST, FLRT3, GGT1, GSN, HIPK2, HMBS, KDM6B, KIAA0100, MINK1, MTCH2, NOLC1, NOS3, PARP1, PIGK, PKM, RBM10, SDC3, TNPO1, UQCRC1, YLPM1* (Figure 4A). The KEGG pathway enrichment analysis revealed that 18 candidate genes might be associated with metabolism, heme biosynthesis, caspase-mediated cleavage of cytoskeletal proteins, and neutrophil degranulation (Figure 4B).

To further explore candidate Se-related pathogenic genes in KD patients, the disease enrichment analysis was performed to identify the relationship between diseases and Se-related genes through the CTD database. The top 20 enriched diseases are shown in Figure 4. Among the top 20 diseases, 2 cardiovascular diseases might be closely related with KD. Then, 7 gene genes were identified, including *ALAD, RBM10, GSN, GGT1, ADD1, PARP1*, and *NOS3*.

To further elucidate the pathogenic genes from 24 candidate Se-related genes, PPI analysis were proposed to prioritize candidate disease genes, which combined with genes from microarray in KD, and genes from proteomics in KD. As shown in Figure 4, top 9 significant genes in accordance with scores and nodes were *NOS3, PARP1, ATRX, GSN, HMBS, PKM, ALAD, GGT1*, and *KDM6B*.

Taking into account the functional role of genes and previous research, *GSN, ADD1*, and *NOS3* are the most likely Se-related genes contributing to the pathogenesis of KD.

### 2.6 Proofreading and validation for the candidate genes

To verify the reliability of these selected genes, we carried out proofreading and validation with UK Biobank, which providing exome-sequence data and 791 phenotypes data from 49,960 participants. Because Keshan disease has a strong correlation with chronic heart failure, all related phenotypes (7) were selected from all the phenotypes (791). For 199 genes from burden analysis, 7 related phenotypes (Data set 1) were further respectively compared with 17 different disease categories (Data set 2) and all the phenotypes (Data set 3) with Fisher’s exact test methods, and the top 20 significant genes were illustrated in Figure 5, including *NEXN, TAF1C, FUT4, ALAD, ZNF608, STX2, SCNN1G, MINK1, YLPM1, ITGA10, MYBPC3, TYW1, GSTZ1, NPHP4, PHLPP1, FANCD2, NMNAT1, COL5A1, AHNAK*, and *DMRTA2*. Remarkably, *NEXN, TAF1C, FUT4, ALAD, ZNF608*, and *STX2* were clustered together and relatively different in 17 different disease categories and all categories. Notably, one DCM-causative gene, *NEXN* was found among these 6 genes.

**Figure 5.**
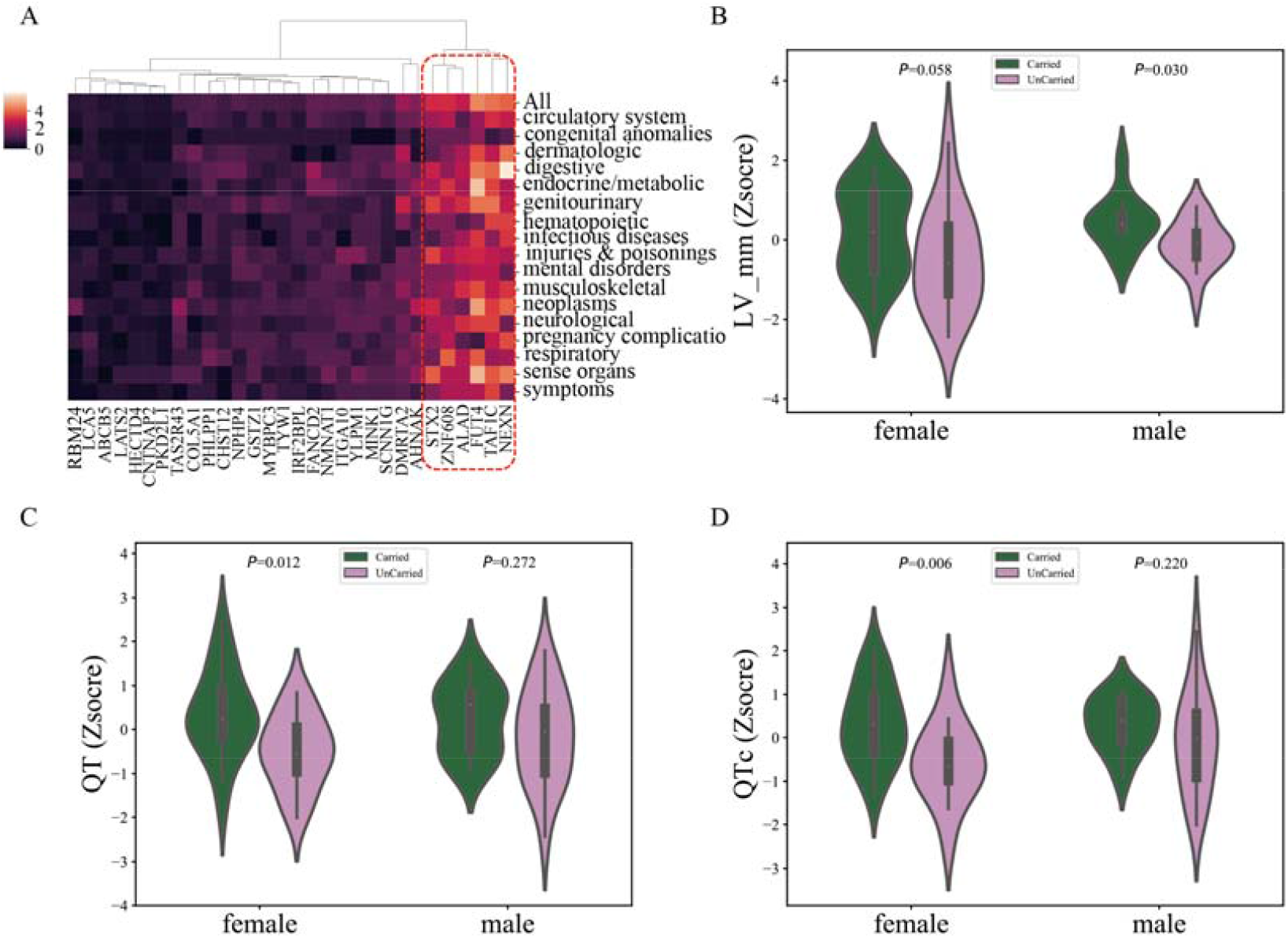
Validation for the candidate genes. (A) The candidate genes from burden analysis were validated with UK Biobank. (B) The left ventricular diameter was analyzed between carried and non-carried mutation patients with the six genes. (C) The left ventricular diameter was analyzed between carried and non-carried mutation patients with the six genes. (D) The QTc was analyzed between carried and non-carried mutation patients with the six genes.

Clinical indicators were further combined to analyze patients with and without the six genes mutations. The left ventricular diameter were significant different between carried mutation and non-carried mutation patients (Figure 5B). The QT and QTc were significant different between carried mutation and non-carried mutation female patients with the six genes, while not in male patients (Figure 5C and 5D).

### 2.7 Proofreading and validation for *ALAD*

To find the most likely pathogenic genes in KD, the candidate genes from four different analyses were taken intersection. Notably, *ALAD* is the only candidate pathogenic gene identified by all analyses, indicating that the gene is highly repetitive and reliable.

We further confirmed the phenotype based on the information of *ALAD* deletion mutants in the mouse gene database MGI (Table 1), and found that the *ALAD* homozygous mutants were lethal before the formation of the atrial septum, indicating that *ALAD* is important in the development of the heart. *ALAD* heterozygous mutants have gender differences in phenotypes, such as reduced grip strength in heterozygous male mice and decreased average red blood cell volume in female mice. This suggesting that *ALAD* might play an important role in the pathogenesis of KD.

**Table 1.**
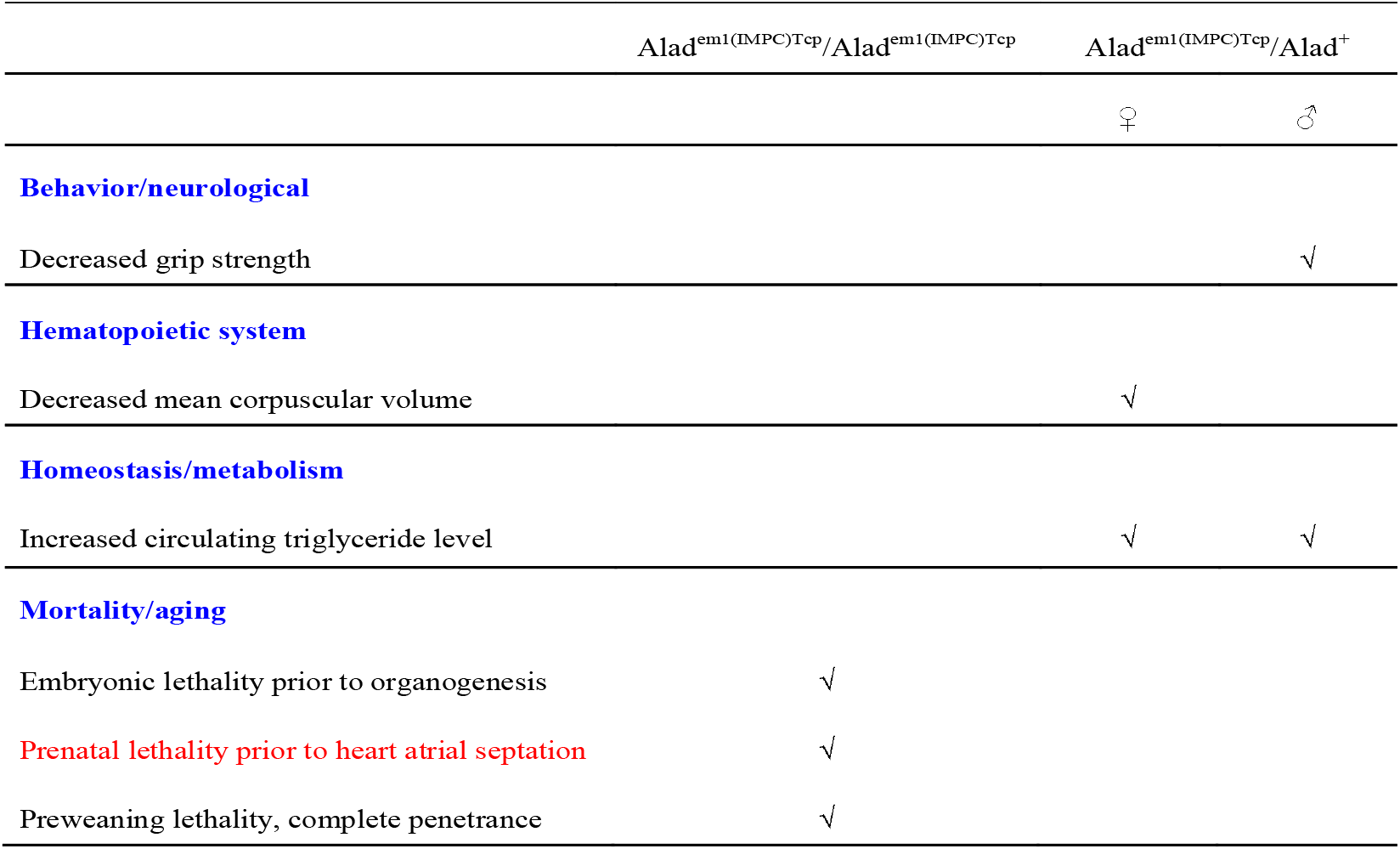
Phenotypic information of *ALAD* deletion mutants in MGI database

## 3 Discussion

Keshan disease is described primarily as dilated cardiomyopathy, manifested as systolic and diastolic dysfunction, heart failure, various arrhythmias and even sudden death [3]. The effective treatments used in KD are not available so far. Fortunately, an in-depth understanding of the molecular mechanisms underlying DCM opens the door to explore Keshan disease [12,13]. As for KD, we identified various DCM-causative genes, encoding for diverse proteins of the sarcomere (*MYH6, MYH7, MYBPC3*, and *MYPN*), cytoskeleton (*TTN, ACTN2, LDB3, SYNM, DMD, FHL2, FKTN, VCL, FLNC, MURC*, and *NEBL*), desmosomes (*DSP, DSC2, DSG2, PKP2*, and *CTNNA3*), nucleus (*RBM20, ANKRD1, PRDM16, ZBTB17, ALPK3*, and *LRRC10*), ion channel (*SCN5A, ABCC9*, and *KCNQ1*), extracellular matrix (*LAMA2*, and *LAMA4*), and mitochondria (*TXNRD2*). There are various these known biological processes, including the sarcomere, cytoskeleton, and ion channel, leading to systolic and diastolic dysfunction in DCM as previous reported. These results suggested that these genes might be involved in the regulation of KD pathogenesis.

Keshan disease tends to be accompanied with conductive arrhythmia, congested heart failure, and cardiogenic shock, in which ion channels may be the important mediators. These three sodium channel genes, *SCN10A, SCN1B*, and *SCNN1G*, were related to each other through PPI analysis. The mutation of *SCN10A*, a sodium channel, was linked to prolonged cardiac conduction in human, and *SCN10A*-dificient mice exhibited a shorter PR interval than controls [15]. Furthermore, *SCN10A* mutation was even found to induce sudden cardiac death [16]. In addition, the mutation of *SCN1B* (encoding the sodium channel β1 subunit), impacted action potential generation in cardiomyocytes [17]. Multiple mutations of *SCN1B* were detected in patients with arrhythmogenic right ventricular cardiomyopathy [18]. *SCNN1G* (encoding epithelial sodium channel γsubunit), were reported to be associated with hypertension [19]. These results suggested that these genes might regulate arrhythmia in KD.

Keshan disease is clinically manifested as systolic and diastolic dysfunction, mainly driven by force-generating mechanism of myocardial contraction and Calcium handling. This seven-myocardial contraction related genes, *VCL, DTNA, NEXN, MYBPC3, GSN, MYOT*, and *PRKAG2*, were found to form an interactive network through PPI analysis. In particular, cytoskeleton-related genes (*DTNA, NEXN*, and *VCL*) and sarcomere-related gene (*MYBPC3*) were previously reported as causative genes in dilated cardiomyopathy. Moreover, the overexpression of Gelsolin (*GSN*), the actin-binding proteins, enhanced cardiomyoblast cell apoptosis [20]. In addition, the mutant Myotilin (*MYOT*), a Z-disc protein, heavily aggregated together with other Z-disc-related proteins [21]. The above evidence indicated that this interactive network might be essential to maintain the function of myocardial contraction in KD patients.

It is thought that immune-inflammatory mediated cardiomyocyte apoptosis, clinically manifested in Keshan disease. Among these nine immunity related genes (*CANX, FUT4, GGT1, HIF1A, INPPL1, JAK2, NOS3, PARP1*, and *TFRC*), the two cores *JAK2* and *TFRC* were found to form an interactive network. The activation of *JAK2* induced apoptosis of myocardial cells under oxidative stress [22]. In addition, the calnexin (*CANX*) was essential for autoimmune in mice [23]. The above evidence suggested that these genes might regulate immune process to influence cardiomyocyte apoptosis. Se deficiency was thought to closely connect with KD, indicating Se-related genes might play a critical role in the process of Keshan disease. Notably, three candidate Se-related genes, Gelsolin (*GSN*), α-adducin (*ADD1*), and nitric oxide synthase 3 (*NOS3*), were determined as most likely pathogenic genes in KD. For example, the overexpression of *GSN* enhanced cell apoptosis and induced cardiomyocyte hypertrophy. KD tends to be accompanied by cardiac hypertrophy and cardiac cell apoptosis, indicating *GSN* might mediate these phenotypes in KD [20]. Moreover, *ADD1*-dificient mice triggered hypertension [24]. In addition, it is identified *NOS3* polymorphisms and an increase in *NOS3* gene expression in DCM, and *NOS3* generates nitric oxide (NO) for vascular smooth muscle relaxation [25]. Thus, although these three genes have not directly related to Keshan disease, the above evidence indicates that these genes may be involved in the occurrence and development of Keshan disease.

The gene *ALAD* encodes δ-aminolevulinic acid dehydratase (ALAD), which is composed of 8 identical subunits and is the key enzyme in the second step of heme synthesis [26]. Hemoglobin is a key protein that carries and transports oxygen in the body with protoporphyrin IX (heme) as a prosthetic group [27]. Its main functions are oxygen transfer, electron transfer, and biosensor. If the activity of ALAD is abnormal, it will hinder the synthesis of hemoglobin and cause a series of diseases [28,29].

It should be pointed out that myoglobin, which is homologous to hemoglobin, only exists in the myocardium and striated muscle, with heme as a prosthetic group, and its biological function is to help myocardial cells transport oxygen to mitochondria [27]. In addition, the hypoxic HIF-1 signaling pathway was extensively enriched, *ALAD* and HIF-1α constitute a protein-protein interaction network, and the ATP biosynthetic pathway was enriched in this study. Furthermore, the *ALAD* homozygous mutants were lethal before the formation of the atrial septum, indicating that *ALAD* is important in the development of the heart, which suggested it might play an important role in the pathogenesis of KD.

Nevertheless, further functional experiments are required to investigate the molecular mechanisms related to these candidate genes. Our study offers gene-wide mutation data that will aid our understanding of KD pathogenesis and be a useful source of drug targets for KD treatment.

## 4. Materials and Methods

### 4.1 Collection of peripheral blood samples

Patients with KD were enrolled in Miren County and Renhe District at Sichuan Province in China in the period from 2016 to 2018. 100 unrelated ethnically matched healthy subjects recruited from the Sichuan Provincial People’s Hospital. Any patients diagnosed with KD patients using defined KD criteria in China (WS/T 210–2011). Cardiac functions of all the endemic area subjects were classified referring to the NYHA (New York heart association) classification method. Whole blood samples from 68 KD patients and 100 normal controls were collected in heparinized vacutainer tubes. Enrolled patients need to sign an informed consent form. The research was approved from the ethics committee of the Sichuan Academy of Medical Sciences and the Sichuan Provincial People’s Hospital.

### 4.2 Whole-exome sequencing, variant selection, and annotation

Briefly, we purified DNA from peripheral blood using the QIAamp DNA Blood Mini Kit (Qiagen, Hilden, Germany). Whole-exome enrichment was performed with SureSelect Human All Exon kit V6 (Agilent Technologies, Santa Clara, CA, USA). Genomic DNA library was carried out to sequence using the llumina’s HiSeq X and NovaSeq system (Illumina, San Diego, CA, USA). The sequenced DNA fragments were aligned with Human Reference Genome (NCBI Build 37) based on the Burrows–Wheeler transform. Removal of duplication, realignment, and recalibration were preformed according to Picard tools (http://picard.sourceforge.net/), GATK (http://www.broadinstitute.org/gsa/wiki/index.php/Home_Page). The calling of single Nucleotide Polymorphism (SNP) and insertion-deletion polymorphisms (indels) was performed using GATK3.7 software. The high-confidence variants were annotated with snpEff (Version 4.2; http://snpeff.sourceforge.net/). In addition, the annotations of all variants were further performed using 1000 Genomes Project data (2014 Oct release, http://www.1000genomes.org), the Exome Aggregation Consortium (ExAC; http://exac.broadinstitute.org), EVS (http://evs.gs.washington.edu/EVS), The ClinVar (http://www.ncbi.nlm.nih.gov/clinvar) database, and Online Mendelian Inheritance in Man (OMIM, http://www.omim.org).

### 4.3 Rare variants of DCM-causative genes

In total, 98 DCM-causative genes were chosen to analyze rare variants in Keshan cases and control subjects[12,13]. These genes were considered as reference genes according to at least one of the following criteria: (1) the dysfunction of these genes led to dilated cardiomyopathy, (2) animal experiments verify the phenotype of dilated cardiomyopathy. The selected DCM-causative genes were shown in Supplementary Table S1.

### 4.4 Gene-based burden analysis and enriched diseases analysis for rare variants

To analyze the aggregate association of rare variants in gene level, we performed gene-based burden analysis to obtain a gene-level mutational profile with unrelated Keshan cases (n = 68) and control subjects (n = 100). Rare variants were defined as ‘deleterious variants’ with MAF < 0.01. The statistical method uses Fisher’s exact test to evaluate gene-based burden analysis. A gene-level mutational profile was calculated according to 1000 Genomes Project data and ExAC with MAF< 0.001, MAF< 0.01, or MAF< 0.05; *p*-value < 0.05. All the significant genes were inputted to the Comparative Toxicogenomics Database (CTD; http://ctdbase.org) to obtain candidate gene lists with cardiovascular enriched diseases. The candidate genes were submitted to Reactome (http://www.reactome.org) to calculate the rich factor, and the top 30 enriched pathways were exhibited with the corrected *p* value. Protein–protein interactions of the candidate genes were further detected by the STRING database (see http://apps.cytoscape.org/apps/stringapp), and visualized it with Cytoscape v2.3 software.

### 4.5 Screening of the Se-related genes in gene-based burden results

Se deficiency was closely linked to the occurrence of Keshan disease; we further input all the genes into CTD database from gene-based burden analysis with MAF< 0.001, MAF< 0.01, or MAF< 0.05; *p*-value < 0.05. Then, Se-related proteins were screened through CTD database. A Venn analysis was further conducted to obtain Se-related pathogenic genes through comparing Se-related proteins with candidate pathogenic genes from enriched diseases analysis.

## Data Availability

The raw/processed data required to reproduce these findings cannot be shared at this time as the data also forms part of an ongoing study.

## Notes

### Funding Statement

National Natural Science Foundation of China, Grant/Award Numbers: 81770379, 81500297, 81470521, 81670290

### Author Declarations

The research was approved from the ethics committee of the Sichuan Academy of Medical Sciences and the Sichuan Provincial People Hospital.

